# Body mass index and birth weight improve polygenic risk score for type 2 diabetes

**DOI:** 10.1101/2021.05.16.21257279

**Authors:** Avigail Moldovan, Yedael Y. Waldman, Nadav Brandes, Michal Linial

## Abstract

One of the major challenges in the post-genomic era is elucidating the genetic basis of human diseases. In recent years, studies have shown that polygenic risk scores (PRS), based on aggregated information from millions of variants across the human genome, can estimate individual risk for common diseases. In practice, the current medical practice still predominantly relies on physiological and clinical indicators to assess personal disease risk. For example, caregivers mark individuals with high body mass index (BMI) as having an increased risk to develop type 2 diabetes (T2D). An important question is whether combining PRS with clinical metrics can increase the power of disease prediction in particular from early life. In this work we examined this question, focusing on T2D. We show that an integrated approach combining adult BMI and PRS achieves considerably better prediction than each of the measures on unrelated Caucasians in the UK Biobank (UKB, n=290,584). Likewise, integrating PRS with self-reports on birth weight (n=172,239) and comparative body size at age ten (n=287,203) also substantially enhance prediction as compared to each of its components. While the integration of PRS with BMI achieved better results as compared to the other measurements, the latter are early-life measurements that can be integrated already at childhood, to allow preemptive intervention for those at high risk to develop T2D. Our integrated approach can be easily generalized to other diseases, with the relevant early-life measurements.

## Introduction

Predicting the risk of an individual to develop a specific disease is a key challenge in clinical decision making [1]. Based on such predictions, individuals can be identified for early intervention to prevent, delay the onset or better manage the disease and its outcome. Understanding the genetic component of the disease can highlight individuals at risk based on their genetic profile. Indeed, with more genetic and phenotypic information available for large cohorts, genome-wide association studies (GWAS) have been used to find genetic variants associated with complex diseases and traits [2–4] Nevertheless, in most GWAS studies, variants that are significantly associated with the disease or trait explained only a small fraction of its presumed genetic heritability component. The shortage of GWAS contribution to complex disease risk has been addressed as the missing heritability problem with various explanations that were presented to address it [5–7]. A likely explanation argues that complex diseases are signified by complex intracellular interactions. However, the many variants that are below significance in GWAS, actually affect the trait, and cumulatively contribute to the phenotype even more than the relatively few statistically significant GWAS variants [8, 9]. In light of this possibility, different studies developed polygenic risk scores (PRS) that consider the accumulative effect of millions of genetic markers to predict the probability of an individual to develop a complex disease [1, 10–13]. In some cases, the PRS methodology was able to highlight individuals with the same risk as individuals with rare monogenic mutations linked to a disease. The greater effect on public health reflects the fact that the PRS-based approach cover many more individuals (up to 20 folds) as compared to rare monogenic mutation carriers [14]. In addition, it was shown that the penetrance of rare monogenic high-risk variants in various diseases is also affected by the polygenic risk background as reflected by PRS [15].

The etiology of common complex diseases is presumed to be a combination of both genetic and environmental factors and the interactions between them [16]. Various physical and clinical measures are often taken to highlight individuals with high risk for diseases, and these measures reflect both genetic and non-genetic factors. For example, high body mass index (BMI), which has both genetic and non-genetic components [17], is a major risk factor for type 2 diabetes (T2D) [18, 19]. Birth weight is yet another example of a physical measure that combines effects from both genetic and environmental factors [20]. However, the direction of the association between birth weight and T2D (low birth weight being a risk or also high birth weight), its scale and whether it is sex-dependent are still not clear [21–25].

In this work we asked whether a combined approach that utilizes both genetic factors (e.g., PRS) and quantitative measures (that have non-genetic components) can improve disease prediction. We evaluated this approach by using both the PRS and physical measurements associated with T2D prevalence (BMI, birth weight and comparative body size at age ten) to predict disease risk, based on the UK Biobank (UKB) cohort [26].

Our results demonstrate that such a combined risk predictor significantly enhances prediction as compared to PRS or each of the underlying measures alone. Importantly, our analysis includes early-life measurements, meaning that individuals at high risk can be identified early in life, leading to more effective intervention.

## Methods

### UK Biobank (UKB) data

The analysis in this work is based on the information available for UKB participants [26]. We focused on Caucasians by limiting the analysis to participants who self-reported themselves as White (being White, British, Irish or any other white background [codes 1, 1001, 1002, 1003, respectively, in Ethnic background, UKB data-field 21000]) and being classified as Caucasians based on their genetic ancestry (Genetic ethnic group, data-field 22006). We also required the individuals to have both genotyping data and information on T2D disease status. Disease classification was based on clinical information provided for UKB participants and encoded by ICD-10 code for T2D (E11.X). Additional phenotypes were used for the analysis: BMI (taken at the UKB Assessment Centre, UKB data field 21001), birth weight (based on self-reporting, UKB data field 20022) and comparative body size at age ten (based on self-reporting, UKB data field 1687). In each of the analyses we focused on individuals with the relevant phenotypic information. To address possible sex differences, the analysis was done separately for males and females. Following the filtering steps, 332,338, 184,288 and 318,260 participants were included in the analysis for BMI, birth weight and body size at age ten, respectively. Finally, we focused on participants evaluated at age 40-70 and removed genetic relatives, by keeping only one representative of each kinship group of related individuals from the same sex (recall that analysis was done separately for each sex). This resulted in sets of 290,584, 172,239 and 287,203 participants for BMI, birth weight and body size at age ten, respectively.

### Polygenic risk score calculation

The PRS of an individual is calculated as the weighted sum of his/her allele values over the set of genotyped markers. This score is based on the genotype of each individual and does not considers sex or age. Therefore, we refer to it as a “raw” PRS. Let *m* be the number of markers used for raw PRS calculation, let *G*_*i*_ be the allelic status of marker *i* in a specific individual (*G*_*i*_ ∈ {0,1,2}), and let *w*_*i*_be the weight of marker *i* (based on the association of the marker with the trait). Raw PRS of that individual is then defined as:

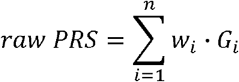

The weights for PRS calculation for T2D on a set of approximately 6.5 million markers (both genotyped and imputed), based on a previous work [14], were downloaded from The Cardiovascular Disease Knowledge Portal (http://www.broadcvdi.org/informational/data). We applied these weights, which had been fit on the UKB data, on the markers of UKB participants to obtain raw PRS values for each individual.

### Composite risk score

In this work, we defined a composite risk score (*CRS*) which is composed of three components: genetic profile (*raw PRS*), phenotypic information *P* (i.e., BMI, birth weight or comparative body size at age 10), and age. For each of the components, we estimate an individual’s disease risk based on the disease prevalence observed within the relevant UKB cohort across individuals with similar scores (e.g., similar raw *PRS* for the genetic component). A weighted sum of the different components is taken to obtain a *CRS* that reflects an individual’s disease risk. The estimated risk scores and weights for each component are learned in a training set and evaluated on a test set (as described below). The rationale behind transforming the original measures into estimated disease prevalence is to allow incorporation of measures that are not necessarily monotonic with respect to disease prevalence. In addition, transforming the measures into disease prevalence also normalizes the different measures, that often span different ranges (e.g., raw PRS and BMI values). The analysis was done separately for each sex.

Formally, we sorted all the individuals in the training set based on their raw *PRS* values and divided them into 100 equal-size bins (i.e., raw *PRS* percentiles of UKB participants). For each bin we calculated T2D prevalence in that bin (i.e., the number of cases divided by the total number of individuals in that bin) and defined it as the genetic risk (*GR*) of the members of that bin. For example, if in a specific bin, 5% of the individuals were reported as having a disease, the *GR* of that bin was defined as 0.05. Thus, the *GR* reflects the actual disease risk in the UKB, based on individuals with similar raw *PRS* scores, sharing the same bin. Let raw *PRS*_*i*_ be the raw *PRS* value of sample. We define *GR* _*i*_ as the *GR* of the bin that raw *PRS*_*i*_ belongs to.

The same procedure was also applied to the phenotypic measure *P*: we sorted all individuals in the training set based on their *P* measures and divided them into 100 equal-size bins and calculated for each bin the phenotypic risk (*PR*) of members of that bin. In the case of comparative body size at age 10, which included only three values (“Thinner”, “About average” and “Plummer”), people were divided to three bins based on this classification and the *PR* was calculated for each of these three predefined bins. We denote the *PR* of individual by *PR*_*i*_.

In addition, we also considered age for the composite score. We divided all individuals in the training set according to their age (measured in rounded years) and for each age calculated the age risk (*AR*) of members with the same age. We denote the *AR* of individual *i* by *AR*_*i*_.

The composite risk score (*CRS*) of sample *i, CRS*_*i*_, was then defined as a weighted sum of the three risk measures:

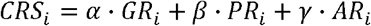

Where:

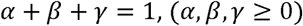

These parameters are trained in and learned in the training set, as described below.

In addition to CRS, we also converted each of the measures alone to disease risk estimates and included age, without including the other measure. Formally, the *PRS* of sample *i, PRS*_*i*_, was defined as follows:

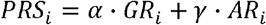

Where:

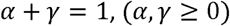

Thus, as opposed to the original raw *PRS, PRS* considers age as well (but does not include the phenotypic measure).

Similarly, for the phenotypic measures BMI and birth weight, we defined a measure risk score that combines them with age, but without PRS. Thus, BMI risk of sample *i* (as opposed to raw BMI that included only the original BMI measurement) was defined as:

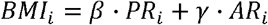

Where:

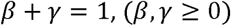

This was also done for birth weight risk (as opposed to raw birth weight) but not to comparative body size at age ten that includes only three distinct values. Finally, we also considered age alone, to examine whether the other measures provide additional predictive information beyond age alone. In that case, it was defined as *AR*_*i*_.

For each measure (CRS, PRS and the phenotypic measures BMI and birth weight), we trained our model on 70% of the individuals (comprising the training set) to estimate the optimal weights (*α, β, γ* depending on the specific measure) that maximize the area under curve (AUC) in the receiver operating characteristic (ROC) for the specific measure. We sampled all combinations for the values of the *α, β, γ* weights, in steps of 0.025 in the range [0,1]. Evaluation of the measures was performed on the remaining 30% of the individuals (comprising the test set), based on odds ratio (OR) analysis, as described below. For the age measure (*AR*) alone there was no weight to learn, but the measure itself (i.e., T2D prevalence per age for each sex) was estimated on the training set and evaluated on the test set.

### Evaluation of the results

We evaluated and compared the different measures (CRS, PRS, BMI, birth weight and age) by examining the resulting T2D OR. For each measure, we divided the participants in the test set into 100 equal-size bins (i.e., percentiles 0-99). We then calculated for each bin its OR. Formally, let *D*_*p*_ be the number of individuals diagnosed with T2D among all individuals in the *p* percentile, and let *H*_*p*_ be the number of individuals not diagnosed with T2D among all individuals in the *p* percentile. Similarly, let *D* _*⌐p*_ and *H* _*⌐p*_ be the number of individuals diagnosed with T2D among all individuals **except** those in the *p* percentile and the number of individuals not diagnosed with T2D among all individuals **except** those in the *p* percentile, respectively. The OR of percentile *p, OR* (*p*) was then defined as:

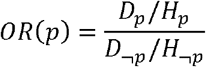

To estimate the robustness of the results (e.g., calculating standard deviations for the OR), we repeated the procedure of randomly dividing the dataset into training and test sets, and evaluating the OR from the classification results of 1000 repetitions.

## Results

### PRS and BMI

In the current study we used the UK Biobank (UKB) cohort [26], focusing on participants whose ethnic background was classified as White, where genotyping information and disease status for T2D was available (see Methods). As there are known sex differences in and T2D prevalence and risk factors [27, 28], we preformed the analysis separately for males and females. Raw PRS (based on [14]), BMI and disease state (case/control) information was available for 290,584 participants, among them 157,813 (54.31%) were females.

**Figure 1A** show the relation between raw PRS and BMI and T2D disease prevalence. As can be seen, both measures were strongly associated with disease prevalence in both sexes. T2D disease prevalence was higher in males as compared to females. The analysis also showed that raw BMI was a better predictor for the disease risk as compared to raw PRS. This was also demonstrated with respect to OR across the different percentiles **(Figure 1B)**. For example, the OR in the 99^th^ percentile was 8.62 vs. 2.87 and 6.79 vs. 2.84 for raw BMI vs. raw PRS in females and males, respectively. The receiver operating characteristic (ROC) curves also confirmed this. The area under the curve (AUC) of the raw BMI measure was larger than the AUC of the raw PRS measure in both sexes: 0.767 vs. 0.626 and 0.721 vs. 0.629 for raw BMI vs. PRS in females and males, respectively **(Figure 1C)**. These results also indicate that the differences between the two measures were larger in females than in males, and that BMI is a better predictor in females than in males for identifying individuals at high risk to develop T2D.

**Figure 1.**
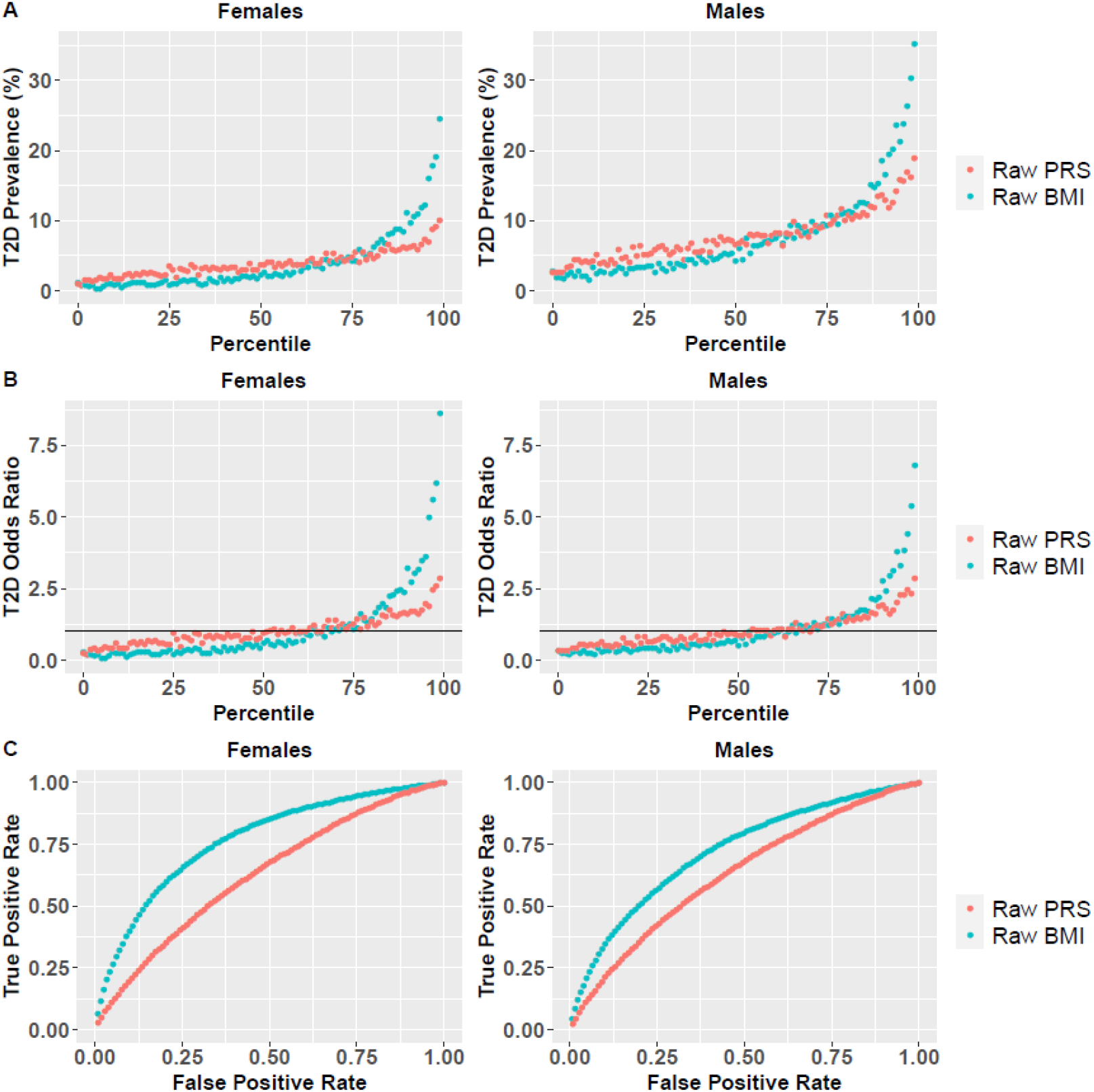
Raw BMI and PRS as predictors for T2D risk. **(A)** T2D disease prevalence based on raw BMI and PRS percentiles for females and males. For each measure (raw BMI and PRS), UKB participants were divided into percentiles, and T2D prevalence was calculated for each percentile. **(B)** T2D odds ratio (OR) for each percentile is shown for females and males, where the horizontal line represents a neutral OR of 1. **(C)** Based on these percentiles, the receiver operating characteristic (ROC) curve is presented for the two measures for females and males, to compare the AUC of the two measures.

Next, we examined whether combining PRS and BMI together can increase their prediction power. For that purpose, we defined a new composite risk score (CRS) which combines both the raw PRS and BMI measures, as well as age. For each of these measures (raw PRS, raw BMI, age) we estimated an individual’s risk based on disease prevalence of people with similar values (e.g., people in the same raw PRS percentile) and combined them into a composite score. The AUC of the combined score was significantly higher as compared to the other measures in both sexes (Wilcoxon signed rank test P-value<10^−16^; **Supplementary Figure S1**). Comparison of OR revealed that for both sexes, BMI exhibited better performance as compared to PRS, but CRS outperformed both measures across all percentiles (**Figure 2**). All measures (BMI, PRS and CRS) outperformed age alone.

**Figure 2.**
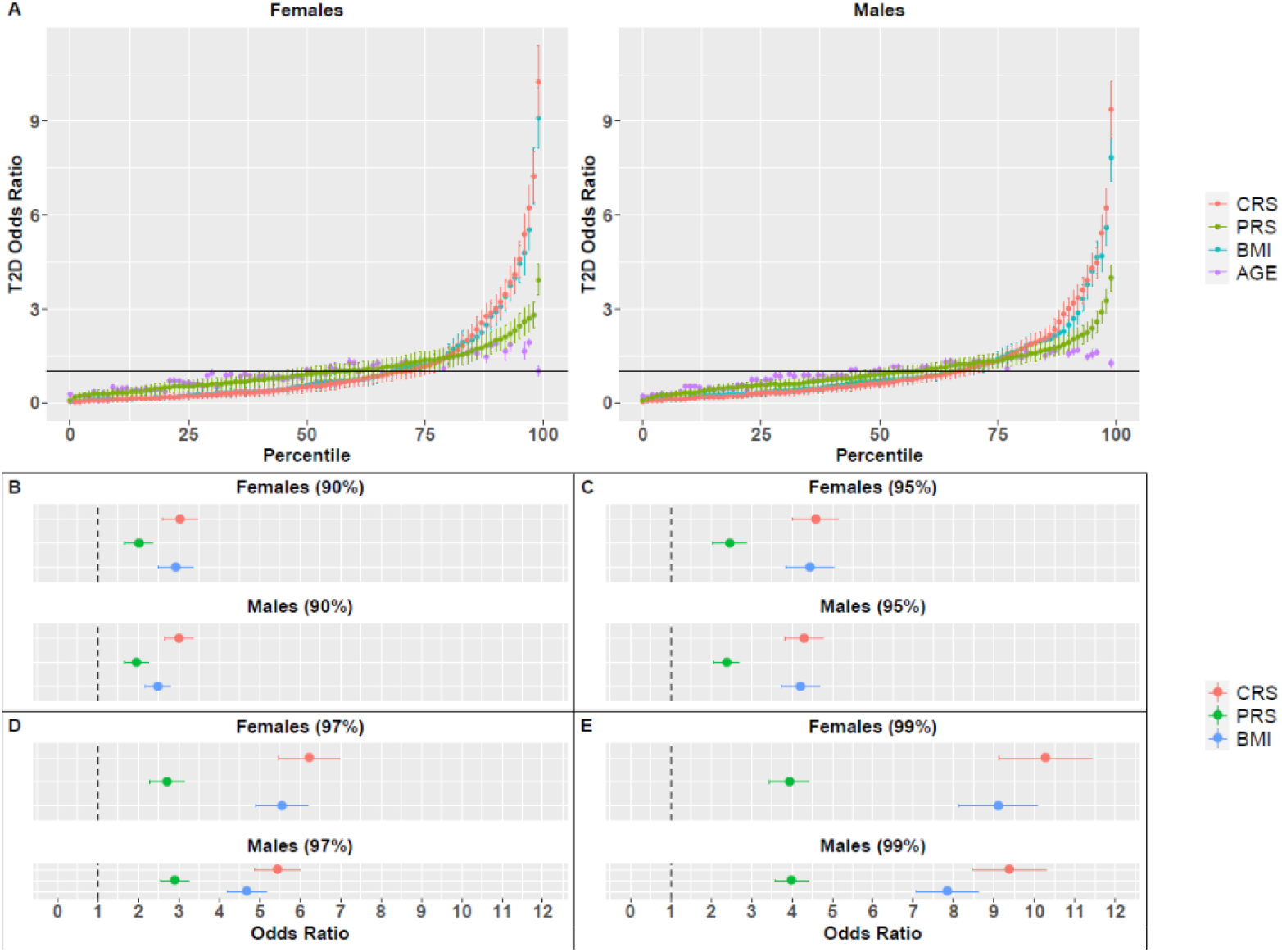
Odds ratio (OR) for T2D, based on BMI, PRS, CRS or age percentiles. **(A)** OR for all percentiles and all measures for females and males. Vertical lines correspond to the standard deviation of the average OR across 1000 random splits of the dataset. The horizontal line represents a neutral OR of 1. OR values for females and males in specific percentiles are also presented: **(B)** 90^th^, **(C)** 95^th^, **(D)** 97^th^ and **(E)** 99^th^.

Specifically, the average OR of the top percentile in males was 3.99, 7.84 and 9.38 for PRS, BMI and CRS, respectively. In females, the average OR of the top percentiles was 3.94, 9.10 and 10.27 for PRS, BMI and CRS, respectively. Additional results of the top percentiles are summarized in **Figures 2C-2E** and **Table 1**. Both PRS and BMI measures that included age achieved higher OR values than the raw PRS and BMI measures that did not include age (**Figure 1B**), demonstrating the importance of adding age into the predictive model.

**Table 1.** Average OR values for T2D for the different measures (BMI, PRS, CRS) by percentiles.

| Sex | Percentile | OR (BMI)<sup>a</sup> | OR (PRS) | OR (CRS) | P-value<sup>b</sup> |
| --- | --- | --- | --- | --- | --- |
| Females | 90 | 2.92±0.43 | 2.01±0.36 | **3.03±0.44** | 3.63×10<sup>-13</sup> |
|  | 95 | 4.44±0.6 | 2.46±0.41 | **4.59±0.57** | <10<sup>-16</sup> |
|  | 97 | 5.54±0.65 | 2.71±0.44 | **6.22±0.75** | <10<sup>-16</sup> |
|  | 99 | 9.10±0.98 | 3.94±0.48 | **10.27±1.16** | <10<sup>-16</sup> |
| Males | 90 | 2.48±0.32 | 1.95±0.29 | **3.00±0.36** | <10<sup>-16</sup> |
|  | 95 | 4.21±0.47 | 2.38±0.31 | **4.30±0.48** | 1.67×10<sup>-12</sup> |
|  | 97 | 4.69±0.49 | 2.90±0.36 | **5.44±0.57** | <10<sup>-16</sup> |
|  | 99 | 7.84±0.76 | 3.99±0.42 | **9.38±0.91** | <10<sup>-16</sup> |

<sup>a</sup>Results include the standard deviation of each measure. Measures with the highest OR for each percentile are bolded. <sup>b</sup>P-value refers to Wilcoxon signed rank test for comparing the OR distributions of the two measures that achieved the average highest OR across 1000 test sets, in each percentile.

These results also demonstrate sex differences with respect to the predictive power of BMI, and therefore of CRS: higher OR values were achieved for females, in accordance with the results reported for the raw measures (**Figure 1**).

### PRS and birth weight

After evaluating BMI, we turned to another physical measure associated with T2D – birth weight. We studied a cohort of 172,239 participants, 105,438 (61.21%) of which were females, who had birth weight values, PRS, and T2D disease state information was available. Similar to the analysis performed for the BMI, we analyzed the association between disease risk and raw birth weight, for males and females separately (**Figure 3**).

**Figure 3.**
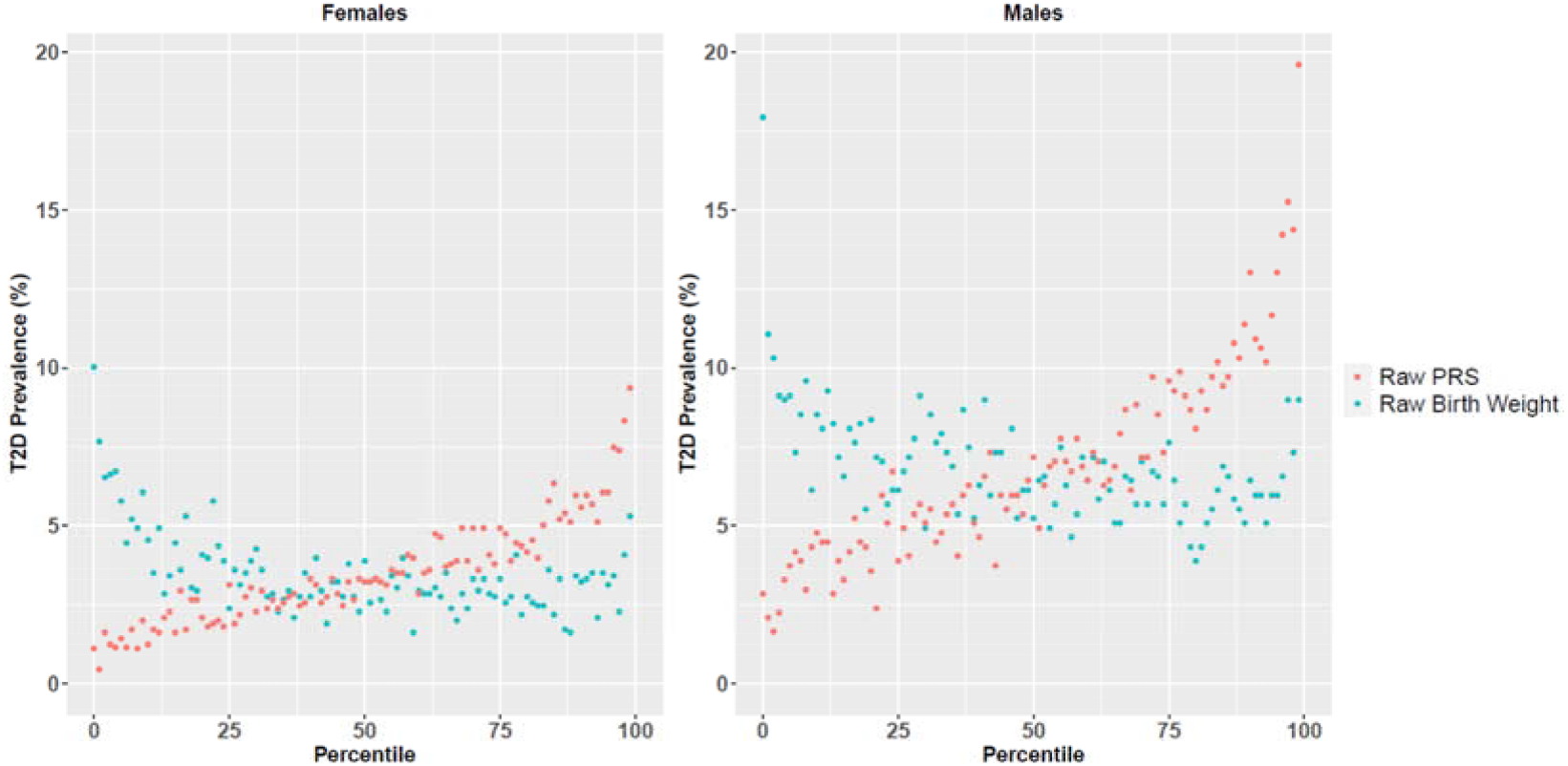
T2D disease prevalence across raw birth weight and PRS percentiles for females and males.

Lower birth weight was associated with higher disease prevalence in both males and females. High birth weight (mainly in the top percentiles) was also associated with higher T2D risk in both sexes, but to a lesser extent.

Next, and similar to the analysis for BMI, we defined a combined score that reflects both the risk associated with birth weight and PRS, based on disease prevalence for different PRS and birth weight percentiles, while also accounting for age. The predictive power (AUC) of the combined score was significantly higher than the individual measures in both sexes (Wilcoxon signed rank test P-value <10^−16^ ; **Supplementary Figure S1**). CRS also achieved higher OR values in the top percentiles (**Table 2**). Specifically, in females it achieved an average OR of 4.64 for the top percentile, compared to 3.81 and 3.62 for PRS and birth weight, respectively. In males the OR values were even higher: 4.83 vs. 4.54 and 3.08 for PRS and birth weight, respectively. For detailed trends across all measurement range (in percentiles) see **Supplementary Figure S2**.

**Table 2.** Average OR values for T2D for the different measures (birth weight, PRS, CRS) by percentiles.

| Sex | Percentile | OR (birth weight)<sup>a</sup> | OR (PRS) | OR (CRS) | P-value<sup>b</sup> |
| --- | --- | --- | --- | --- | --- |
| Females | 90 | 1.84±0.42 | 1.94±0.47 | **2.00±0.45** | 2.25×10<sup>-6</sup> |
|  | 95 | 1.99±0.44 | 2.55±0.53 | **2.59±0.52** | 0.014 |
| | 97 | 2.26±0.48 | 2.78±0.54 | **3.11±0.60** | $<10^{-16}$ |
| | 99 | 3.62±0.57 | 3.81±0.63 | **4.64±0.67** | $<10^{-16}$ |
| Males | 90 | 1.67±0.38 | **1.99±0.44** | 1.97±0.40 | 0.11 |
|  | 95 | 1.83±0.40 | **2.56±0.50** | 2.51±0.50 | 3.39×10<sup>-4</sup> |
| | 97 | 1.94±0.39 | 2.59±0.47 | **2.81±0.51** | $<10^{-16}$ |
| | 99 | 3.08±0.51 | 4.54±0.73 | **4.83±0.72** | $<10^{-16}$ |

<sup>a</sup>Results include the standard deviation of each measure. Measures with the highest OR for each percentile are bolded. <sup>b</sup>P-value refers to Wilcoxon signed rank test for comparing the OR distributions of the two measures that achieved the average highest OR across 1000 test sets, in each percentile.

While BMI was more predictive of T2D risk than birth weight, the latter also significantly improved prediction power (as part of the combined score) over PRS. Comparing males and females, we observed that males had higher OR values in the higher percentiles, for both PRS and CRS measures (but not for birth weight).

### PRS and body size at age ten

Studies have shown that childhood obesity increases the risk for adult T2D and coronary artery disease (CAD) [29, 30]. Information on childhood BMI was not available for UKB participants but a related childhood measure of a comparative body size at age ten was available for 287,203 participants, among them 156,307 (54.42%) were females. While this measure is subjective and retrospective, and included only three predetermined categorical values (thinner, about average and plumper), it was still associated with T2D risk in adulthood (**Figure 4**).

**Figure 4.**
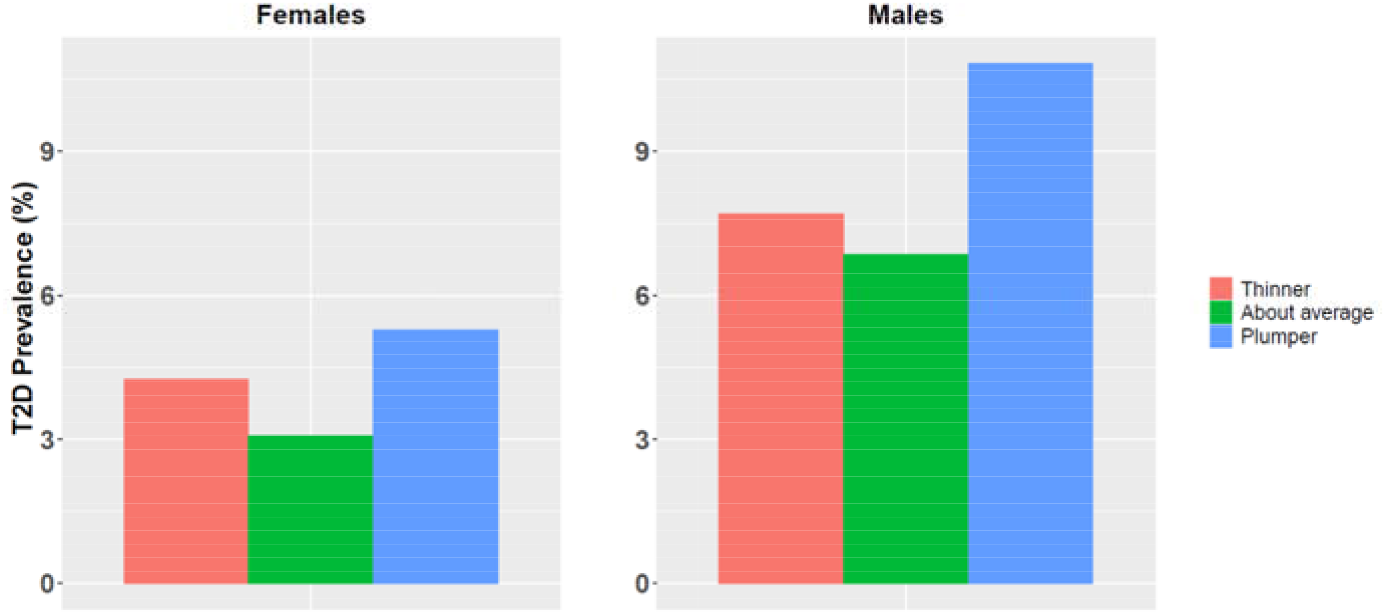
T2D disease prevalence for different categories of body size at age ten for females and males.

People who had described themselves as being plumper at age ten were at higher risk to develop T2D in adulthood compared to people reporting average weight at that age. Similarly, but to a lesser extent, people who described themselves as being thinner at age ten were also at higher risk to develop T2D later in life. This was observed in both sexes These differences in T2D prevalence between the three groups were highly significant (Chi square test P-value<10^−16^).

Next, we defined a combined score that considers PRS, comparative body size at age ten and age. Even with this subjective and simplistic categorical measure, the CRS significantly outperformed PRS with respect to AUC (Wilcoxon signed rank test P-value<10^−16^; **Supplementary Figure S1**) and OR (**Figure 5** and **Table 3**). Results for males and females were very similar, with slightly higher OR values in males (for both PRS and CRS). Specifically, the average OR in the top CRS percentile was 4.18 vs. 3.83 for PRS in females and 4.24 vs. 3.98 in males.

**Table 3.** Average OR values for T2D for PRS and CRS by percentiles.

| Sex | Percentile | OR (PRS)<sup>a</sup> | OR (CRS) | P-value<sup>b</sup> |
| --- | --- | --- | --- | --- |
| Females | 90 | 1.97±0.35 | 2.02±0.36 | 7.71×10<sup>-5</sup> |
|  | 95 | 2.43±0.41 | 2.40±0.38 | 0.031 |
|  | 97 | 2.75±0.43 | 3.00±0.45 | <10<sup>-16</sup> |
|  | 99 | 3.83±0.46 | 4.18±0.49 | <10<sup>-16</sup> |
| Males | 90 | 1.96±0.28 | 2.06±0.29 | <10<sup>-16</sup> |
|  | 95 | 2.38±0.32 | 2.48±0.33 | <10<sup>-16</sup> |
|  | **97** | 2.93±0.35 | **3.07±0.37** | <10<sup>-16</sup> |
|  | **99** | 3.98±0.42 | **4.24±0.42** | <10<sup>-16</sup> |

<sup>a</sup>Results include the standard deviation of each measure. Measures with the highest OR for each percentile are bolded. <sup>b</sup>P-value refers to Wilcoxon signed rank test for comparing the OR distributions of the two measures that achieved the average highest OR across 1000 test sets, in each percentile.

**Figure 5.**
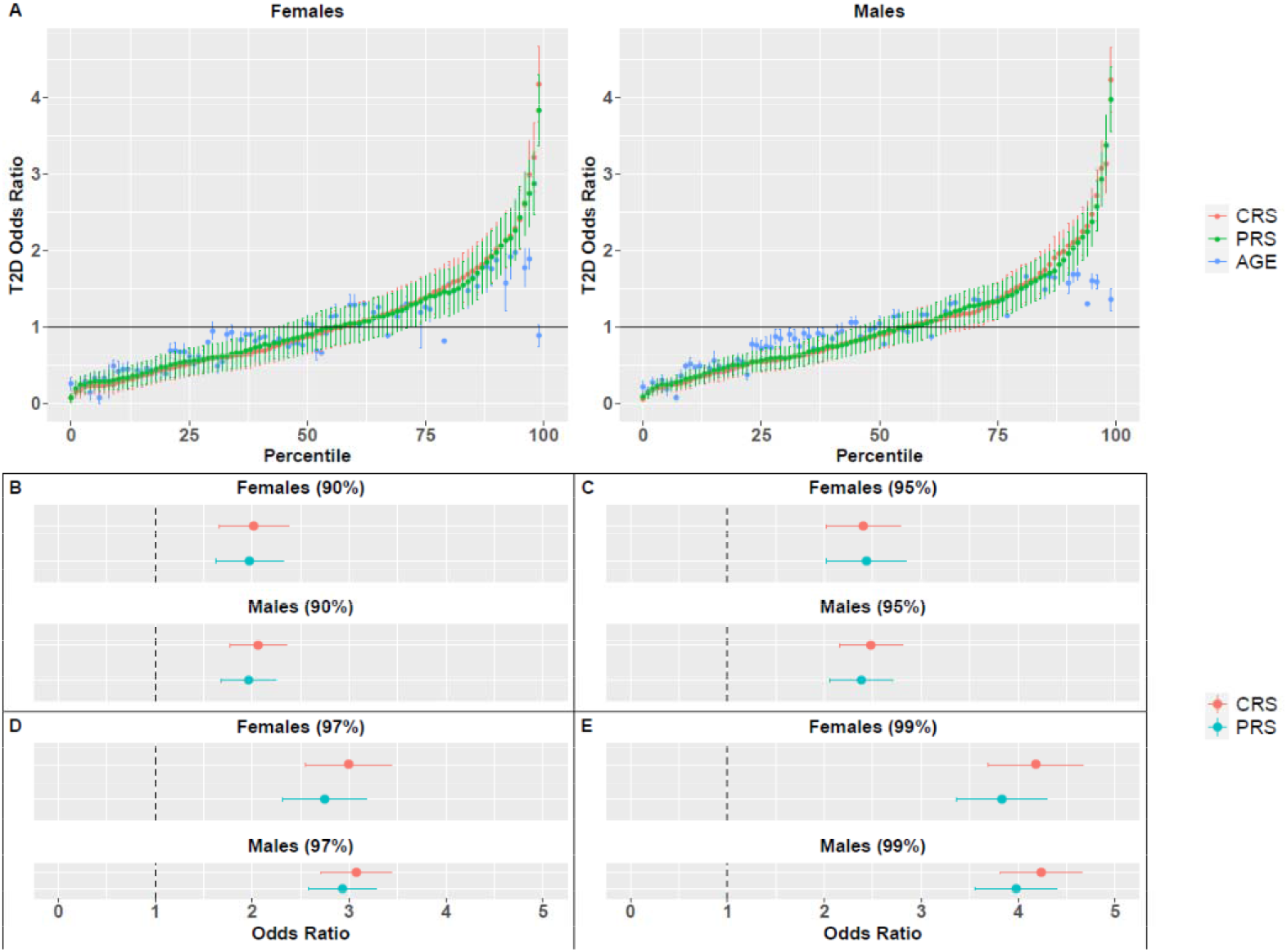
Odds ratio (OR) for T2D, based on PRS, CRS or age percentiles. (A) OR for all percentiles and all measures for females and males. Vertical lines correspond to the standard deviation of the average OR across 1000 random splits of the dataset. The horizontal line represents a neutral OR of 1. OR values for females and males in specific percentiles are also presented: **(B)** 90^th^, **(C)** 95^th^, **(D)** 97^th^ and **(E)** 99^th^.

## Discussion

In recent years, PRS has attracted increasing attention as a potential tool to estimate disease risk for common conditions and diseases based on the genetics of individuals [1, 12]. In the current work we enhanced PRS prediction potential by integrating the raw genetic signal with available physical measures that capture non-genetic (environmental) components of human diseases, focusing on T2D. First, we integrated information on BMI into the PRS model, as high BMI is a well-known risk factor for T2D [18, 19]. We found that while both PRS and BMI can highlight individuals with higher risk to develop T2D, a combined approach was superior to each of the measures alone, for both males and females, demonstrating the added value in such an approach.

Recently, several studies used integrated approaches for disease risk estimation by adding PRS information to standard clinical predictors. Conceptually, these studies applied the combined approach from both sides of its components: either to augment standard disease risk predictors with PRS or to augment PRS with disease risk predictors. Studies that focused on coronary artery disease (CAD) showed no [31] or little [32] improvement when adding PRS to clinically accepted risk predictors. These results raised again the question and the ongoing debate regarding the clinical utility of PRS [1, 33, 34].

A different study on CAD did find significant improvement by adding PRS to the routinely used risk predictors [35]. Another study on CAD, T2D, atrial fibrillation, breast and prostate cancer found that PRS improved the prediction power of such predictors [36]. Similarly, augmenting PRS with additional information such as BMI, and lab results such as HDL and LDL measures improved prediction power for T2D [37]. Similarly, augmenting PRS by traditional measures for cardiovascular disease risk modestly enhanced its prediction power [38]. In addition, a recent study added mortality risk factors to disease PRS to mark individuals with higher mortality risk [39].

Importantly, these studies used measures collected at adulthood while PRS values can be calculated earlier at life to indicate individuals at risk. Indeed, measurements that are taken at adulthood are likely to have stronger prediction power, as more relevant information on the disease and its risk predictors is revealed. However, interventions at the adult stage may be less effective, as some of the biological processes leading to diseases may have already started. Naturally, a composite score that includes adult BMI measures also suffers from this limit. Therefore, we examined whether augmenting PRS with early-life measures can increase their predictive utility. While genetic risk itself cannot be modified, additional risk factors that impact long-term health outcomes and are obtained at early life can be addressed through routine healthcare policy. In our study we used two such early-life measures that were available for many UKB participants: birth weight and three categories of body size at age ten. Similar to previous studies, we found association with low birth rate and high T2D prevalence, with stronger association in females. This is in accordance with the developmental origins theory, which suggests that low birth weight reflects under nutrition in utero that can lead to permanent changes in body functions, posing higher risk for certain metabolic diseases [40]. A weaker association was also found for high birth weight. Importantly, the number of UKB participants that were included in this analysis was relatively large as compared to many previous studies analyzing the relation between birth weight and T2D [22]. A combined approach that included birth weight and PRS improved the prediction power of each of its components. Turning to comparative body size at age ten, we found that adding this information to PRS improved its prediction power as well. Indeed, BMI had a better predictive power as compared to these early-life measures. However, these measures may only partially reveal the component they intend to reflect. Specifically, the body size categories at age ten measure is retrospective, subjective and included only three categories. Therefore, the labels for the body size at age ten only roughly estimated the actual body size at that age. Despite these limitations, early-life measures significantly improved PRS prediction power. We anticipate that more accurate and relevant measures such as childhood BMI or other relevant measures (that are routinely collected at the clinic), as well as their trajectories (across different ages), will further improve disease risk estimation and may inform early intervention.

This work also introduces a revised approach with respect to integrate age and sex into a predictive risk model. Traditionally, the sex of an individual is considered a covariate that is controlled for when learning raw PRS weights [41]. Therefore, when these weights are used, the resulting PRS is no longer affected by sex, and an individual’s PRS is determined solely based on their genetic background, regardless of their sex. In practice, like in other diseases, there are substantial sex differences in T2D prevalence and pathophysiology [27, 28]. In this work we addressed this issue by performing the analysis for each sex separately. Therefore, two people with the same raw PRS value but different sex may be given a completely different risk score. Indeed, we observed differences between the sexes. First, T2D prevalence was much higher in males as compared to females. In addition, T2D risk in the top percentiles for the PRS measure was slightly higher in males. This may perhaps explain why T2D risk in the top percentiles for the CRS measure (which is partially based on the PRS measure) was also higher in males when PRS was integrated with birth weight and comparative body size at age ten. However, when PRS was combined with BMI, the CRS measure achieved higher OR scores in females. This is likely because BMI, which outperforms PRS in its prediction power, is a better predictor in females for highlighting individuals at higher risk for T2D [42].

Similar to sex, age is also often considered a covariate that is controlled for when learning PRS weights. The inferred PRS of an individual is constant and does not change with age. However, similar to other diseases, T2D prevalence increases with age [43]. Here we addressed the role of age as a principal risk factor by adding it into the predictive model. As a result, our score reflects an individual’s risk to develop T2D around their age, and it changes throughout life, resulting in risk score trajectories.

We designed our combined risk score to be simple and easy for application and generalization. Thus, the PRS measure was based on raw PRS weights that had been calculated in a previous work [14]. While we focused on T2D, such summary statistics are available for numerous other diseases and traits (e.g., the Polygenic Score Catalog, [44]). Therefore, with additional relevant phenotypes and measures (based on the nature of the disease), our approach can also be applied to other complex diseases. In addition, we converted each of the measures used in the study into disease prevalence measures (based on the average disease prevalence in people with similar values of that measure). This conversion allowed us to easily integrate measures whose relation with disease prevalence is not monotonic (e.g., birth weight and comparative body size at age ten), and to integrate measures of different scales without explicit normalization. We integrated the different measures through a simple linear model. Taken together, this method can be applied relatively easily to various diseases, using various relevant measurements.

Even with this simplified approach, we achieved significant improvements that highlighted the importance of an integrated approach to estimate disease risk. Future works can further improve this through complementary ways to calculate and integrate risk factors. Below we briefly outline some suggestions for such improvements, mainly in the integration of sex and age into the model. First, our sex-specific approach was applied after the calculation of the raw PRS values, which can also be calculated for each sex alone. Indeed, several recent works used sex-specific PRS values because of the putative role of sex in many diseases and mortality [39, 45]. Second, for simplicity of the combined approach, age was taken as an independent measure with a constant effect. However, the role of some T2D risk factors changes throughout life [46]. Specifically, the weight of the genetic component of T2D varies across different ages of onsets [47], and this can lead to differential power of PRS to predict disease risk across different age groups, as was demonstrated in other diseases [48, 49]. Hence, the integration of age into the model can be done in more sophisticated ways (e.g., nonlinear), reflecting the apparent different weights of each component at different ages.

In summary, we demonstrated the benefit of adding measures to enhance PRS prediction. Specifically, we integrated PRS with early-life measures to pave the way for early intervention. We hope this will encourage future work on the integration of PRS with additional measures to provide more accurate clinical risk estimates for T2D and other complex diseases.

## Data Availability

The entire analysis is based on UKBB aggregated data.

## Acknowledgments

We thank Ido Margaliot for useful discussion. We thank Center for Interdisciplinary Data Science (CIDR) and the CSE system team for support in data storage.

## Ethics and Regulation

The UK-Biobank application ID 26664 (Linial lab). Ethical committee approval, The Hebrew University #13082019.

## Supplementary Figures

**Supplementary Figure S1.**
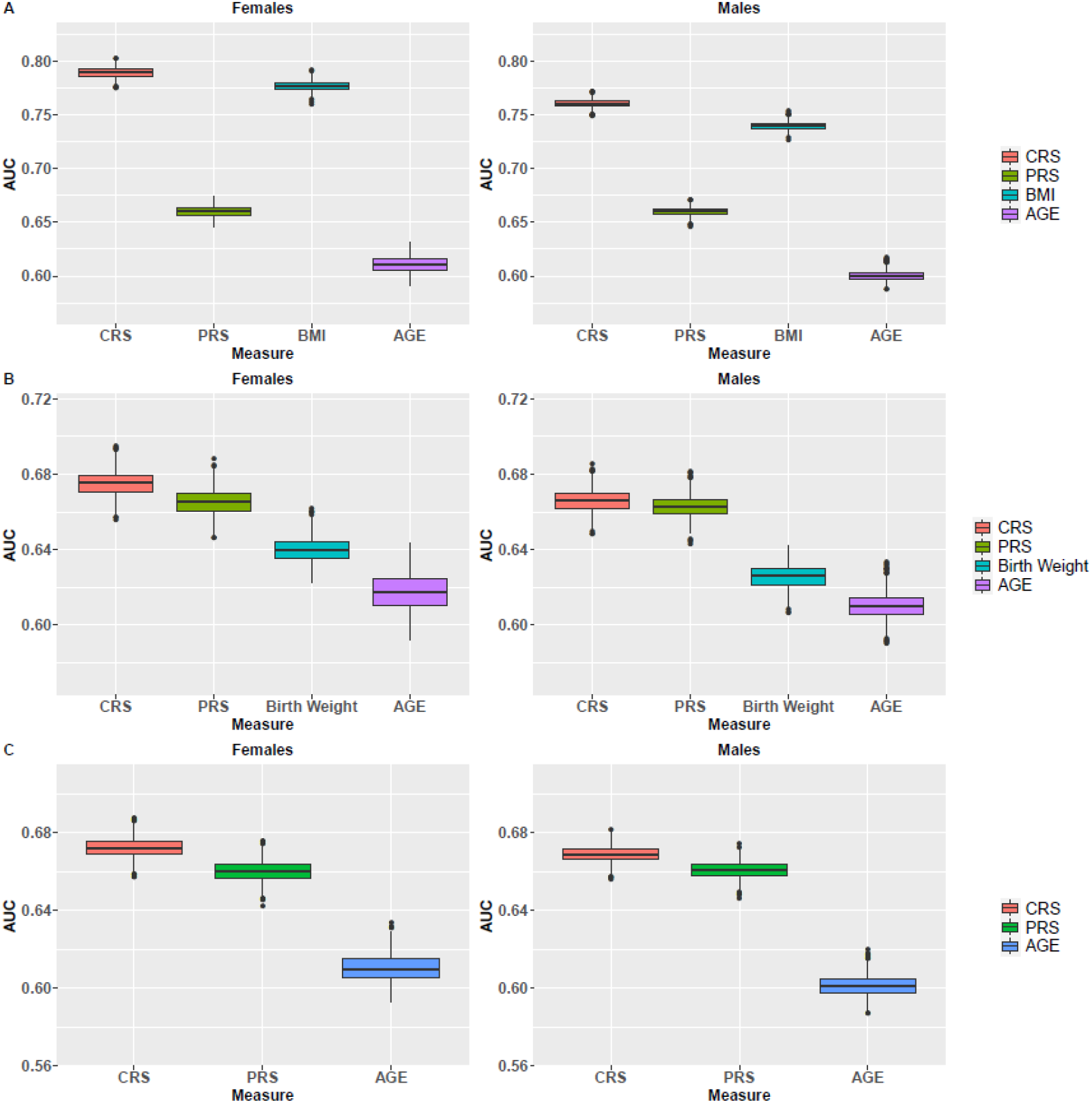
Comparison of AUC values in the test sets for the different measures. In each analysis (BMI, birth weight and comparative body size at age ten), we compared the AUC values of the different measures across 1000 random test sets. Results are shown for the analysis of PRS with **(A)** BMI, **(B)** birth weight and **(C)** comparative body size at age ten. In all cases, the AUC of the combined measure (CRS) achieved significantly higher values as compared to the AUC of all other measures (Wilcoxon signed rank test P-value<10^−16^).

**Supplementary Figure S2.**
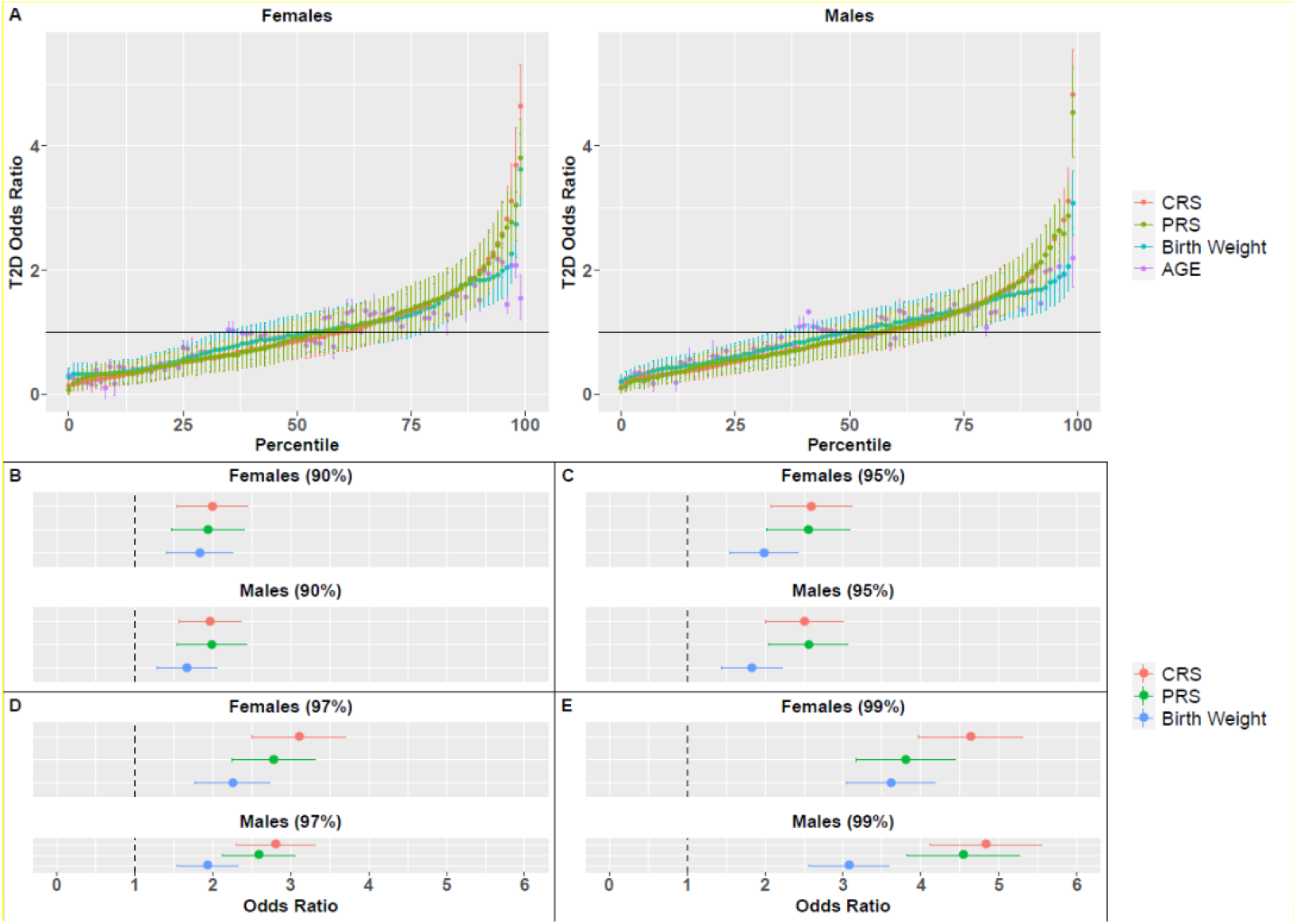
Odds ratio (OR) for T2D, based on birth weight, PRS, CRS or age percentiles. **(A)** OR for all percentiles and all measures for females and males. Vertical lines correspond to the standard deviation of the average OR across 1000 random splits of the dataset. The horizontal line represents a neutral OR of 1. OR values for females and males in specific percentiles are also presented: **(B)** 90^th^, **(C)** 95^th^, **(D)** 97^th^ and **(E)** 99^th^.

